# The cost-effectiveness of common strategies for the prevention of transmission of SARS-CoV-2: a flexible policy online platform

**DOI:** 10.1101/2020.08.13.20166975

**Authors:** Zafar Zafari, Lee Goldman, Katia Kovrizhkin, Peter Muennig

## Abstract

**Background:** The added value of interventions to prevent the transmission of SARS-CoV-2 among university affiliates is uncertain but needed as universities attempt to remain open.

**Methods:** We use a decision-analytic simulation to examine the cost-effectiveness of common interventions to reduce SARS-CoV-2 transmission. We use Columbia University for reference values but our approach centers around an online model that allows users to tailor the model and interventions to their local conditions and existing strategies. All interventions are compared relative to implementing the Centers for Disease Control and Prevention (CDC) guidelines alone. Results. At prevalence rate of actively infectious cases of COVID-19 in the community surrounding the university of 0.1%, using a symptom-checking mobile application is cost-saving relative to CDC guidelines alone and the university can expect to remain open. At a prevalence of 1%, standardizing masks will be cost-saving. At a prevalence rate of 2%, thermal imaging cameras cost $965,070 (95% credible interval [CrI] = $198,821, $2.15 million)/quality-adjusted life year (QALY) gained. One-time testing on entry costs $1.08 million (95% CrI = $170,703, $3.33 million)/QALY gained. Weekly testing costs $820,119 (95% CrI = $452,673, $1.68 million)/QALY gained. Upgrades to ventilation systems or installation of far-ultraviolet C lighting systems will be cost-effective at a willingness-to-pay threshold of $200,000/QALY gained only if aerosols account for 86-90% of all on-campus transmission of SARS-CoV-2.

**Conclusions:** The value of interventions to prevent transmission of SARS-CoV-2 vary greatly with the prevalence rate of actively infectious cases of COVID-19 in the community surrounding the university.

**Key Points:** Universities are struggling to remain open in the face of COVID-19 infections among affiliates. There are a number of modalities to reduce the transmission of SARS-CoV-2, but it is unclear which are worthwhile investments for any given university.

## Introduction

In September of 2020, roughly half of U.S. universities and colleges allowed at least some students back for in-person instruction. ^1-4^ Re-opening protocols for universities in the US are largely based on a core set of principles set by the Centers for Disease Control and Prevention (CDC), ^5^ including social distancing, facial coverings, an emphasis on handwashing, and enhanced cleaning procedures. ^5^ The CDC indicates that testing university affiliates for COVID-19 has not been systematically studied. ^5^ Many additional screening and preventive measures are available, but the value of implementing them has not been assessed.^1,6^

To address such uncertainties, we developed the Columbia Covid-19 Model. ^7^ This is a user-accessible model that allows different universities to alter input parameters via an online interface. Universities can use our model to tailor their strategies based upon local conditions and existing interventions. ^7^

## Methods

### Overview

The Columbia Covid-19 Model is decision-analytic model that deploys a Monte Carlo simulation. In this model, a cohort of students and a cohort of staff/faculty cycle daily through a 91-day semester. ^7^ As each day passes, it calculates the risk of an event (e.g., an infection, hospitalization, or death) based on the prevalence of actively infectious cases of COVID-19 in the community where a university is situated.

For the present analysis, we used Columbia University as a case study because we have extensive information on the socio-demographic characteristics of university affiliates, novel survey data, and cost data on an array of preventive investments. Our model adopts a societal perspective that allows for comparisons across cost-effectiveness analyses using the Consolidated Health Economic Evaluation Reporting Standards (CHEERS) guidelines. ^8^

### Interventions

We compared CDC guidelines (social distancing, protective measures, and maintaining a healthy environment) to a “do nothing” status quo. ^9^ We then compared the CDC guidelines with a requirement that all university affiliates self-report COVD-19associated symptoms. We chose to do so *a priori* because virtually all universities in the US had, at a minimum, implemented the CDC guidelines, so they represent the “status quo.” ^1^ Our online model also allows for comparisons against no intervention at all. ^7^

We then assessed a policy that universities provide standardized, high quality masks.^13,14^ This reduces the number of students using thin, loosely-fitting masks that may be less effective than snug two-ply masks.^13,14^ (See **Table 1** for assumptions and **Table 2** for input values for this paper.)

**Table 1.**
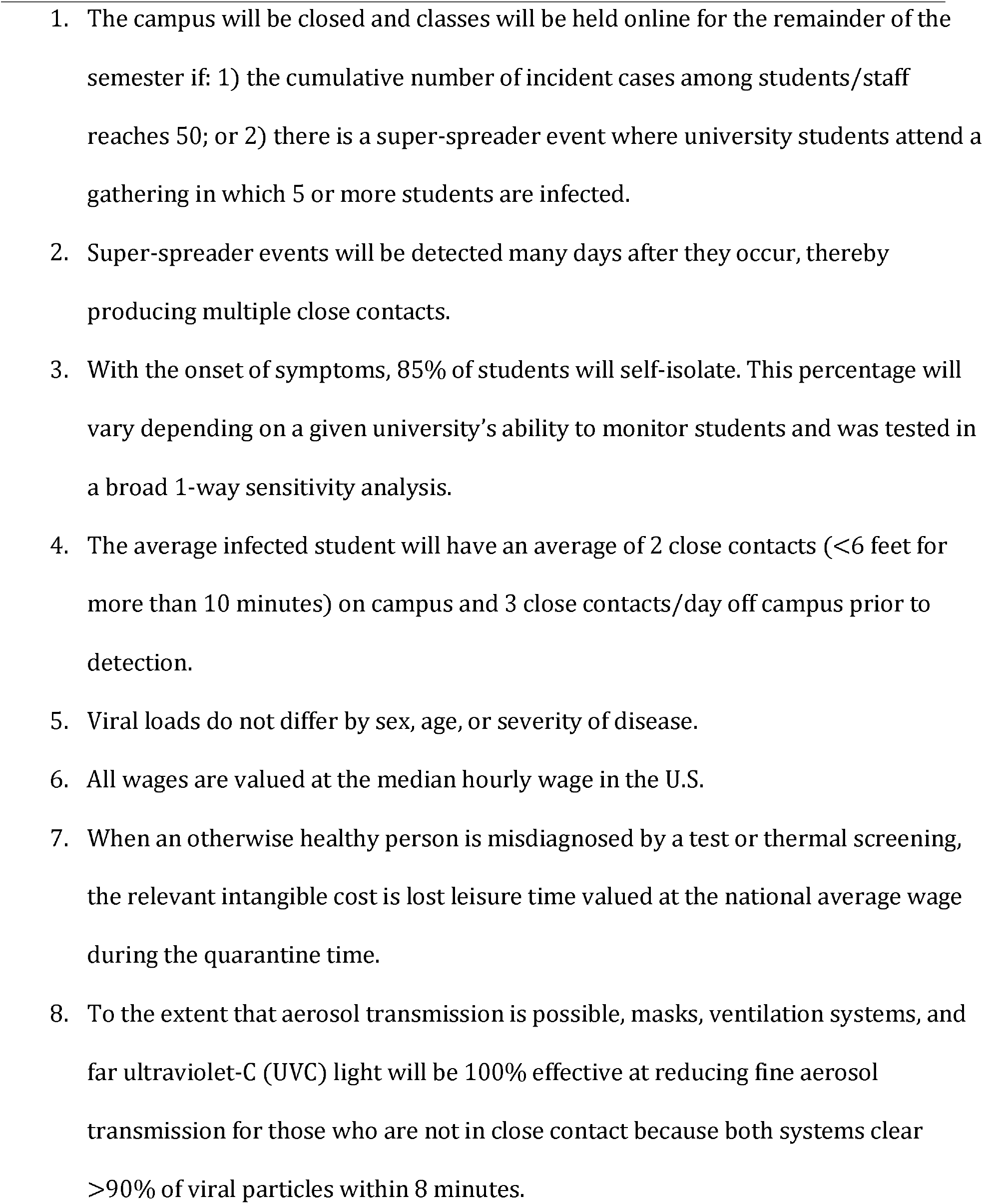
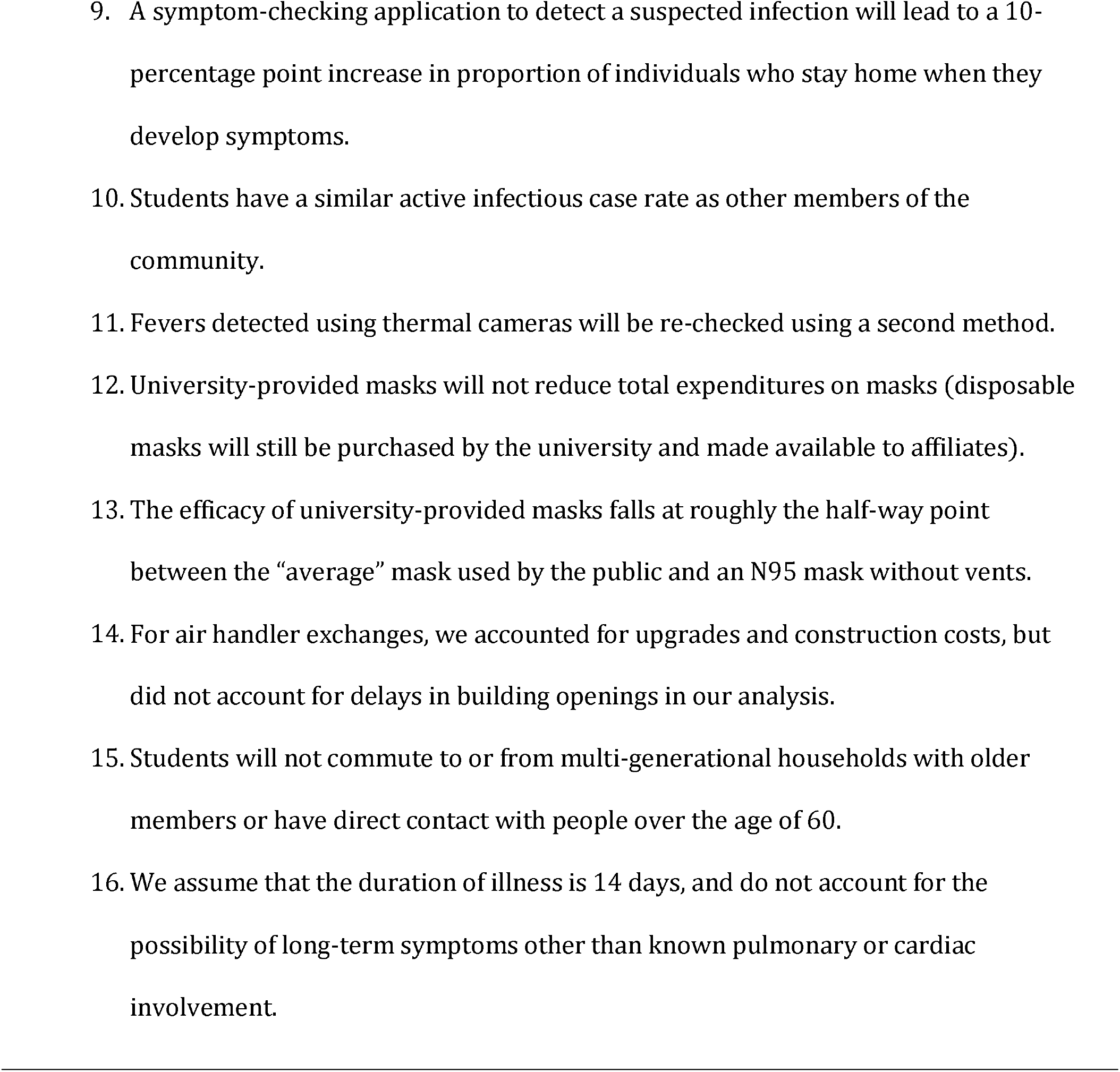
Major assumptions used in modeling the cost-effectiveness of strategies to improve infection control for Covid-19 in the university setting.

**Table 2.**
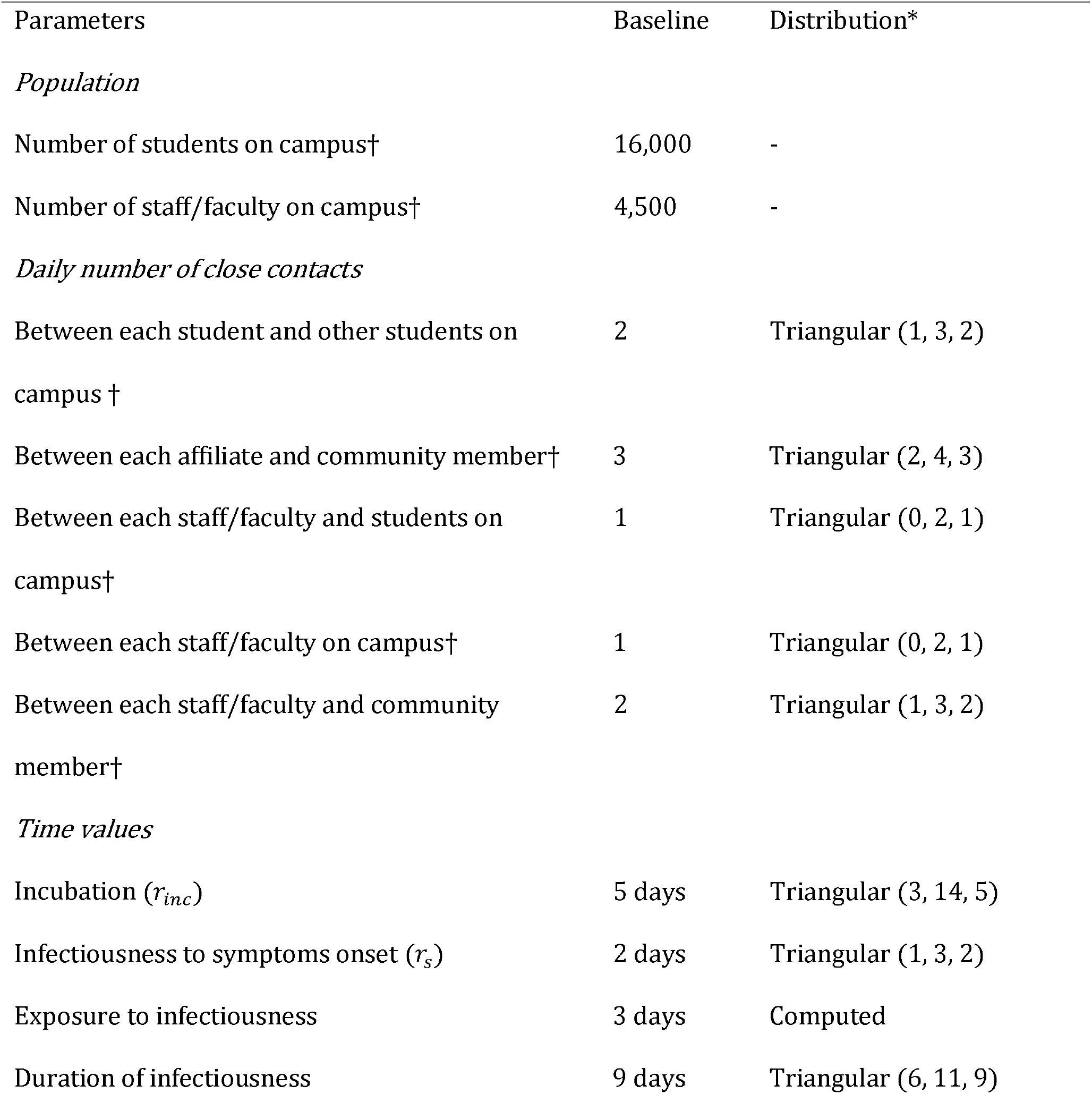
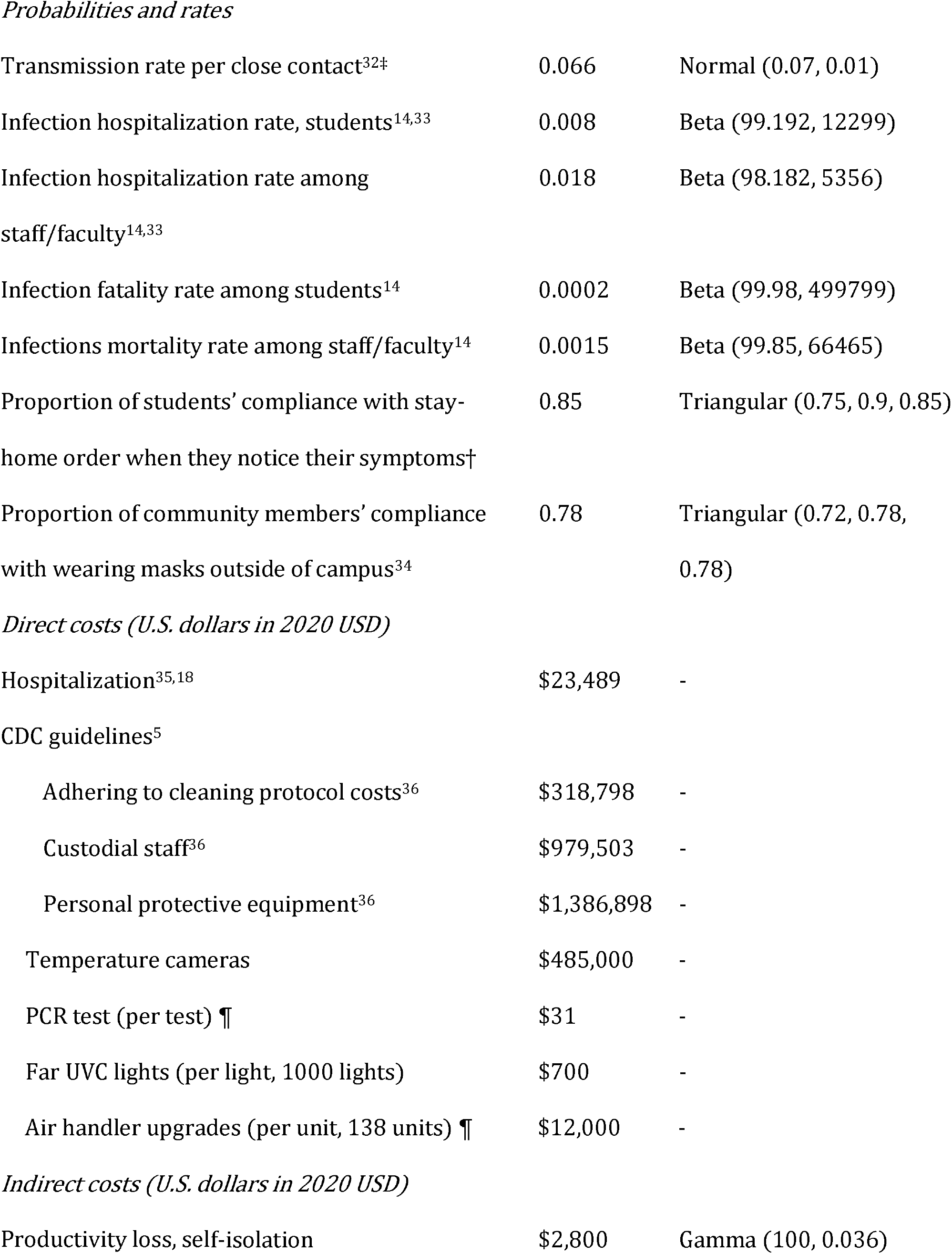
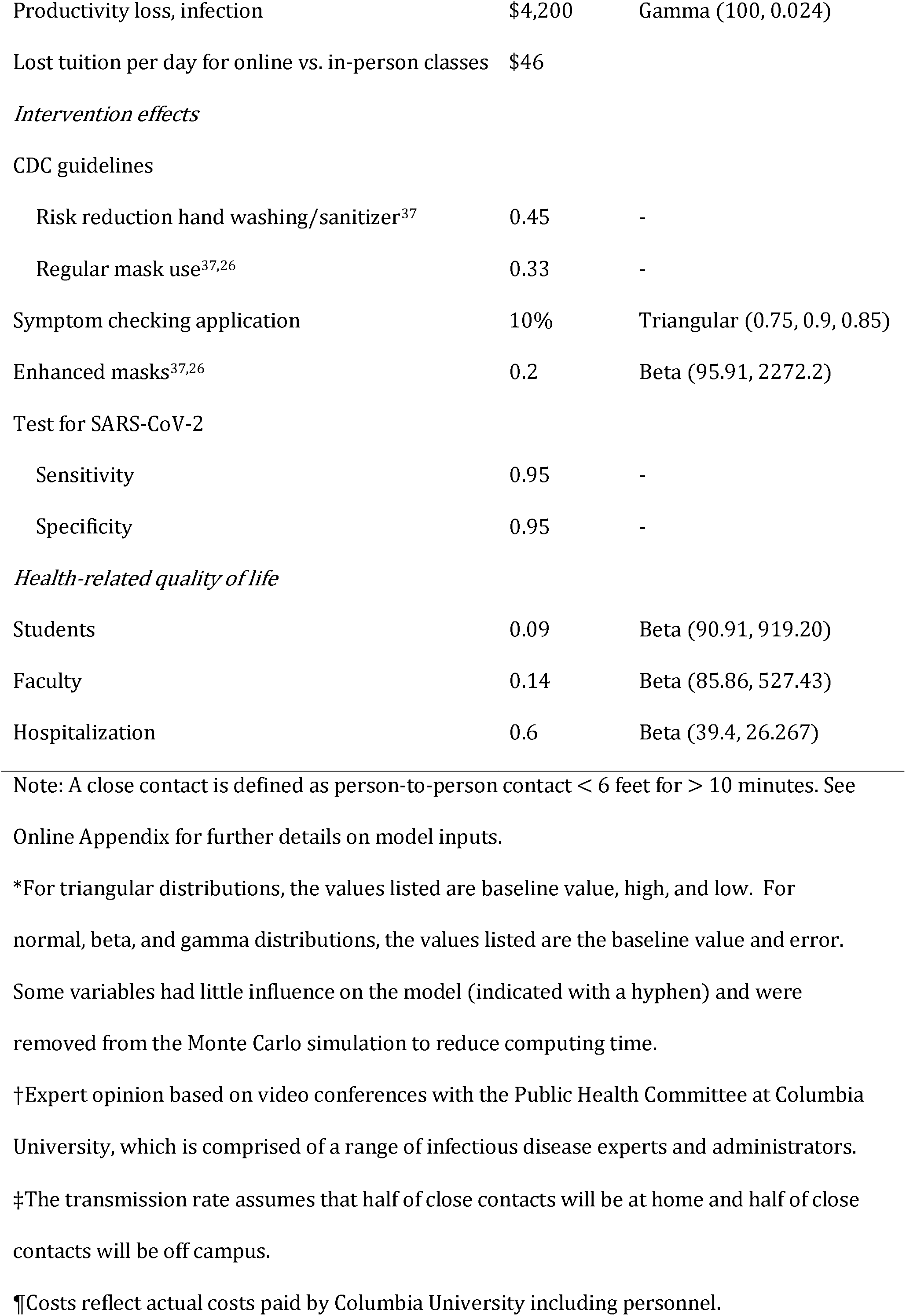
Total costs and probabilities used as model inputs for estimating the cost-effectiveness of strategies to improve infection control for Covid-19 in a university setting with 16,000 students and 4,500 employees on campus during a 90-day semester.

| Parameters | Baseline | Distribution* |
| --- | --- | --- |
| *Population* |  |  |
| Number of students on campus† | 16,000 | - |
| Number of staff/faculty on campus† | 4,500 | - |
| *Daily number of close contacts* |  |  |
| Between each student and other students on campus † | 2 | Triangular (1, 3, 2) |
| Between each affiliate and community member† | 3 | Triangular (2, 4, 3) |
| Between each staff/faculty and students on campus† | 1 | Triangular (0, 2, 1) |
| Between each staff/faculty on campus† | 1 | Triangular (0, 2, 1) |
| Between each staff/faculty and community member† | 2 | Triangular (1, 3, 2) |
| *Time values* |  |  |
| Incubation ($r_{inc}$) | 5 days | Triangular (3, 14, 5) |
| Infectiousness to symptoms onset ($r_s$) | 2 days | Triangular (1, 3, 2) |
| Exposure to infectiousness | 3 days | Computed |
| Duration of infectiousness | 9 days | Triangular (6, 11, 9) |
| *Probabilities and rates* |  |  |
| Transmission rate per close contact<sup>32‡</sup> | 0.066 | Normal (0.07, 0.01) |
| Infection hospitalization rate, students<sup>14,33</sup> | 0.008 | Beta (99.192, 12299) |
| Infection hospitalization rate among staff/faculty<sup>14,33</sup> | 0.018 | Beta (98.182, 5356) |
| Infection fatality rate among students<sup>14</sup> | 0.0002 | Beta (99.98, 499799) |
| Infections mortality rate among staff/faculty<sup>14</sup> | 0.0015 | Beta (99.85, 66465) |
| Proportion of students' compliance with stay-home order when they notice their symptoms† | 0.85 | Triangular (0.75, 0.9, 0.85) |
| Proportion of community members' compliance with wearing masks outside of campus<sup>34</sup> | 0.78 | Triangular (0.72, 0.78, 0.78) |
| *Direct costs (U.S. dollars in 2020 USD)* |  |  |
| Hospitalization<sup>35,18</sup> | \$23,489 | - |
| CDC guidelines<sup>5</sup> |  |  |
| Adhering to cleaning protocol costs<sup>36</sup> | \$318,798 | - |
| Custodial staff<sup>36</sup> | \$979,503 | - |
| Personal protective equipment<sup>36</sup> | \$1,386,898 | - |
| Temperature cameras | \$485,000 | - |
| PCR test (per test) ¶ | \$31 | - |
| Far UVC lights (per light, 1000 lights) | \$700 | - |
| Air handler upgrades (per unit, 138 units) ¶ | \$12,000 | - |
| *Indirect costs (U.S. dollars in 2020 USD)* |  |  |
| Productivity loss, self-isolation | \$2,800 | Gamma (100, 0.036) |
| Productivity loss, infection | \$4,200 | Gamma (100, 0.024) |
| Lost tuition per day for online vs. in-person classes | \$46 | |
| *Intervention effects* |  |  |
| CDC guidelines |  |  |
| Risk reduction hand washing/sanitizer<sup>37</sup> | 0.45 | - |
| Regular mask use<sup>37,26</sup> | 0.33 | - |
| Symptom checking application | 10% | Triangular (0.75, 0.9, 0.85) |
| Enhanced masks<sup>37,26</sup> | 0.2 | Beta (95.91, 2272.2) |
| Test for SARS-CoV-2 |  |  |
| Sensitivity | 0.95 | - |
| Specificity | 0.95 | - |
| *Health-related quality of life* |  |  |
| Students | 0.09 | Beta (90.91, 919.20) |
| Faculty | 0.14 | Beta (85.86, 527.43) |
| Hospitalization | 0.6 | Beta (39.4, 26.267) |

Note: A close contact is defined as person-to-person contact < 6 feet for > 10 minutes. See Online Appendix for further details on model inputs.
\*For triangular distributions, the values listed are baseline value, high, and low. For normal, beta, and gamma distributions, the values listed are the baseline value and error. Some variables had little influence on the model (indicated with a hyphen) and were removed from the Monte Carlo simulation to reduce computing time.
†Expert opinion based on video conferences with the Public Health Committee at Columbia University, which is comprised of a range of infectious disease experts and administrators.
‡The transmission rate assumes that half of close contacts will be at home and half of close contacts will be off campus.
¶Costs reflect actual costs paid by Columbia University including personnel.

Next, we assessed temperature monitoring cameras at facility entry points^11, 12^ to prevent entry of people with a fever. This strategy is commonly used, and has the effect of reducing the number of students and employees on campus (thereby increasing social distancing) even in the absence of COVID-19.

Finally, we assessed one-time entry (“gateway”) testing for SARS-CoV-2 for all affiliates ^10^ as well as weekly testing for SARS-CoV-2.

Because many universities are experimenting with improving airflow, we also conducted one-way sensitivity analyses on methods to remove particulate aerosols (ventilation systems with minimum efficiency reporting value [MERV]-13 filters or far ultraviolet C [far-UVC] light).^11^ In our study, we measure the impact of replacing existing air handlers entirely and removing re-circulation systems (the process that Columba University used). At the time of writing, the proportion of COVID-19 cases attributed to aerosol transmission was unknown, and a one-way sensitivity analysis allows the reader to assess whether the proportionate reduction is plausible.

All of the interventions vary by adherence to mask use. We therefore present the results of our analysis at different levels of mask use.

### Outcome measures

We examined: 1) the incremental cost of each intervention after accounting for medical and intangible costs (e.g., in-person versus online classes); 2) the incremental quality-adjusted life years (QALYs) gained,^8,12^ where QALYs capture both longevity and health-related quality of life. A QALY is scaled between zero, representing death, and 1.0, representing one year of life spent in a perfect health; and 3) the incremental cost-effectiveness ratio (ICER, changes in costs divided by the changes in QALYs).

### Model specification

Students and staff/faculty were treated as two separate but interacting populations with different baseline ages, average number of close contacts, exposures, and risks of illness, hospitalization, and death due to Covid-19. ^13^ We obtained data from Columbia University on the age of each student, staff, and faculty member. Age-specific risks of hospitalization and death were obtained from the literature and from the CDC,^14,15^ and a weighted risk was calculated separately for students and staff/faculty based upon the age distribution of each.

We divided the simulation cohort into four mutually exclusive states: susceptible (those who have not developed the disease and are at risk), infected (those who are currently infected and contagious), recovered (those who were infected in the past but are currently recovered), and death. The cycle length of the model is one day, and the time horizon is over the semester (91 days).

We assessed risk using Mathematica’s 19andMe model. ^13^ This model accounts for the daily probability of becoming infected among susceptible population based on the average number of close contacts, the transmission rate per close contact, and the estimated prevalence of infectious cases inside and outside of the campus. For each susceptible student and staff/faculty, the probability of becoming infected outside of campus was calculated as follows:

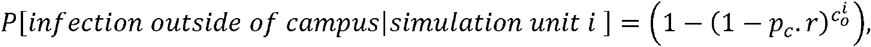

where *p_c_* represents the prevalence of infectious Covid-19 cases in local community outside of campus; *r* is the transmission rate per a close contact; and 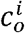 represents the average number of daily close contacts that each simulation unit i (students or. staff/faculty) makes in the local community outside of campus.

The prevalence of actively infectious cases in the surrounding community, defined as the number of people infected within the past 7 days ^16^ per 100,000 people after correcting for underreporting. ^17^

To account for the proportion of population wearing face masks outside of campus, we assigned a multiplier factor, 1−*C_o_.RR_wearing mask_*_;_ where *C_o_* represents the compliance rate with wearing face masks in local community outside of campus; and *RR_wearing mask_* represents a risk reduction associated with wearing face masks.

Similarly, the probability of becoming infected inside the campus was calculated as follows:

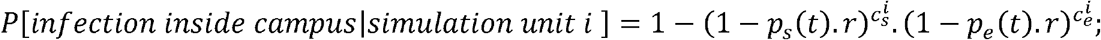

where *p_s_*(*t*) and *p_e_*(*t*) represents the prevalence of infectious cases among students, and staff/faculty, respectively, at time *t*; *r* is the transmission rate per a close contact; and 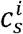, and 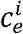 represent the average number of close contacts that each simulation unit *i* makes with students and staff/faculty, on campus, respectively.

Once infected, three consecutive phases of disease progression are possible, denoted as the time between: 1) the primary exposure and infectiousness; 2) infectiousness and onset of symptoms (during which they can infect a susceptible individual but they do not manifest symptoms and therefore do not self-quarantine); and 3) symptom onset until the end of infectiousness. For asymptomatic infected individuals, the model excludes the second phase. At the end of the third phase, infected individuals are classified as ‘recovered.’ Infected affiliates are exposed to a chance of illness, hospitalization, and death.

In addition, infected affiliates incur risk-adjusted costs and lost QALYs associated with having and treating Covid-19 infection. ^18^ Lost productivity and leisure time were valued at the average American wage. ^19^ Intangible costs associated with online versus in-person instruction were valued using a survey administered to students who had experienced learning in each format. Risk tolerance was assessed using a standard gamble exercise (see Online Appendix).

To evaluate the effects of different interventions, two types of risk reductions were modeled: 1) the removal of infected affiliates from the university community via screening/tests; and 2) a reduction in SARS-CoV-2 transmission while on campus (by applying the adjusted odds ratio of infection associated with different interventions).

We assumed that the campus would close and instruction would switch to online-only learning for the remainder of the semester under two conditions: 1) if the model reached a total of 500 cases of COVID-19 cases among students/staff/faculty (based upon New York State requirements for closure); or 2) if a super-spreader event (defined as 5 or more university affiliates from a given campus infected on a single day) occurred. We single out super-spreader events because they can produce multiple asymptomatic infections at once, and are usually only uncovered during extended contact investigation (if at all). As such, they can lead to a spike in infections on campus that overwhelm the university’s ability to close and clean different areas of the campus.

We modeled the probability of a super-spreader event based upon the prevalence of infectious cases of disease in the community, *p_c_*, a standard gamble-based risk assessment administered to students that revealed students’ preferences for participation in community parties, *p_party_* as the daily probability of students’ participation in a community party, and the average number of attendees in a community party, *N_party_*, as follows:

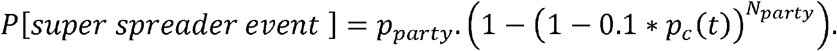

Because roughly 10% of exposed cases are, on average, responsible for 80% of subsequent infections, we multiplied *p_c_* by 0.1 to approximate the prevalence of super spreaders in a community party.

See the Online Appendix for additional methods, outcome measures, and full list of model parameters. ^7^

### Analyses

We ran Monte Carlo simulations on all variables simultaneously and 1-way sensitivity analyses on variables that produced a large influence on the ICER. We assessed 3 scenarios of the prevalence of actively infectious cases of COVID-19 in July, 2020: New York City, “low prevalence” (roughly 0.1%); Texas, “moderate prevalence” (1%); and Florida, “high prevalence” (2%). ^17^ Results are reported as mean and a 95% “credible interval” based upon a random sampling of values from multiple distributions (see **Table 2**). Our model was built on the R statistical platform (The R Foundation, Inc). We deployed R-Shiny upon R, an interface that allows users to alter model parameters. ^7^

## Results

### No guidelines in place

At a 0.1% prevalence of actively infectious cases (similar to New York City in July, 2020) ^21^, roughly 350 out of the 20,500 university affiliates we model would contract COVID-19 infections over the 91-day semester if the university were opened with no CDC guidelines in place (“status quo”). However, if the prevalence of actively infectious cases were 1% (Texas in July, 2020) or 2% (Florida in July, 2020), these numbers would rise to 2420 and 4340 infections, respectively.

### CDC guidelines in place

At a community prevalence of 0.1%, implementing CDC recommendations would reduce the number of infections to roughly 230, and the university would remain open the entire semester at a cost of $2.93 million/QALY gained (95% CrI = $1.23 million/QALY gained, 11.89 million/QALY gained).

At a community prevalence of 1%, implementing the CDC guidelines would likely be cost-saving (**Table 3**), but the “status quo” projections of infections would never be reached because the university would have to shut down after 36 days (95% CrI = 26, 29 days). At a prevalence of 2%, implementing CDC guidelines is less favorable than at a 1% prevalence ($65,657/QALY gained; 95% CrI = savings, $2.30 million/QALY gained) because the university would likely close after 18 days (95% CrI = 13-25 days).

**Table 3.** Average number of days that the university will remain open, incremental costs, incremental quality-adjusted life years (QALYs), and incremental cost-effectiveness ratio (ICER) for each intervention relative to guidelines from the Centers for Disease Control and Prevention (social distancing, mask use, sanitization of spaces).

|  | Days<br>university<br>open | Incremental cost | Incremental<br>effectiveness | ICER |
| --- | --- | --- | --- | --- |
| **100 Cases/100,000** |  |  |  |  |
| Check symptoms | 91 (91, 91) | -\$13 (-\$27, -\$4) | 0.019 (0.005, 0.043) | Cost-saving |
| Gateway testing | 91 (91, 91) | \$2.60m (\$2.32m, \$2.90) | 0.064 (0.034, 0.11) | \$40.9m (\$24.6m, \$78m) |
| Weekly testing | 91 (91, 91) | \$29.2m (\$26.2m, \$32.6m) | 0.48 (0.28, 0.78) | \$60.7m (\$37.5m, \$105m) |
| 2-ply mask | 91 (91, 91) | \$164k (\$164k, \$164k) | 0.11 (0.027, 0.27) | \$1.44m (\$597k, \$6.13m) |
| Thermal imaging | 91 (91, 91) | \$3.86m (\$3.38m, \$4.40m) | 0.066 (0.039, 0.11) | \$58.9m (\$35.7m, \$102m) |
| **1000 Cases/100,000** |  |  |  |  |
| Check symptoms | 36 (26, 49) | -\$107k (-\$739k, -\$36) | 0.057 (0.014, 0.23) | Cost-saving |
| Gateway testing | 37 (27, 50) | \$1.29m (\$339k, \$2.07m) | 0.67 (0.31, 1.20) | \$1.94m (\$302k, \$6.1m) |
| Weekly testing | 42 (31, 58) | \$10.7m (\$7.15m, \$15.2m) | 4.26 (2.52, 6.31) | \$2.52m (\$1.28m, \$5.22m) |
| 2-ply mask | 37 (27, 50) | -\$780k (-\$2.04m, \$164k) | 0.48 (0.094, 1.17) | Cost-saving |
| Thermal imaging | 36 (26, 50) | \$1.33m (\$708k, \$2.08m) | 0.56 (0.34, 0.86) | \$2.36m (\$921k, \$5.63m) |
| **2000 Cases/100,000** |  |  |  |  |
| Check symptoms | 18 (13, 25) | -\$32.6k (-\$721k, -\$50) | 0.035 (0.008, 0.27) | Cost-saving |
| Gateway testing | 20 (15, 27) | \$1.27m (\$311k, \$2.02m) | 1.18 (0.55, 2.07) | \$1.08m (\$171k, \$3.33m) |
| Weekly testing | 22 (16, 30) | \$5.88m (\$4.13m, \$8.55m) | 7.18 (4.43, 10.2) | \$820k (\$453k, \$1.68m) |
| 2-ply mask | 19 (14, 26) | -\$335k (-\$1.29m, \$164k) | 0.46 (0.086, 1.18) | Dominant* |
| Thermal imaging | 18 (13, 26) | \$915k (\$247k, \$1.35m) | 0.95 (0.58, 1.40) | \$965k (\$199k, \$2.15m) |

#### 0.1% prevalence

At a 0.1% community prevalence rate of active infectious cases, a symptom-checking application would be cost-saving and produce 0.02 QALYs gained (95% CrI = 0.01 QALYs gained-0.04 QALYs gained) relative to implementing the CDC guidelines alone (**Table 3**). University-provided 2-ply masks, temperature monitoring cameras, gateway testing, and weekly testing would each cost >$1 million dollars/QALY gained relative to CDC recommendations (**Table 3**). For example, weekly testing would cost $60.70 million (95% CrI = $37.5 million, $104.5 million)/QALY gained.

#### 1% prevalence

At a 1% community prevalence rate of infectious cases, university-provided masks would likely become cost-saving (95% CrI = cost saving, $1.74 million/QALY gained), but, all other interventions would cost >$1.9 million dollars/QALY gained.

#### 2% prevalence

At a 2% community prevalence rate of infectious cases, thermal imaging cameras cost $965,070 (95% CrI = $198,82, $2.15 million)/QALY gained. Gateway testing would cost $1.08 million (95% CrI = $170,703, $3.33 million)/QALY gained, and weekly testing would cost $820,119 (95% CrI = $452,673, $1.68 million)/QALY gained.

#### Sensitivity analyses

At prevalence rates of 0.3% or higher, early university closure is possible. Therefore, the perceived value of in-person instruction becomes an important variable. If students are willing to pay 12% of the regular tuition (rather than our baseline estimate of 48%) for online classes, gateway testing becomes cost-effective at a willingness-to-pay threshold of $200,000/QALY gained. At this prevalence rate, university-provided 2-ply masks and implementing the CDC guidelines become cost-saving.

Weekly testing is sensitive both to the number of close contacts per student on campus and the transmission rate. However, even as the number of close contacts per student increases, the value is unlikely to reach the willingness-to-pay threshold of $200,000/QALY gained at a prevalence of 0.1% (see Online Appendix). When the prevalence rate reaches 2%, however, weekly testing is a strategy that is cost-saving when the number of contacts or the transmission rate is higher than those we use in the baseline scenario (**Figure 1**).

**Figure 1.**
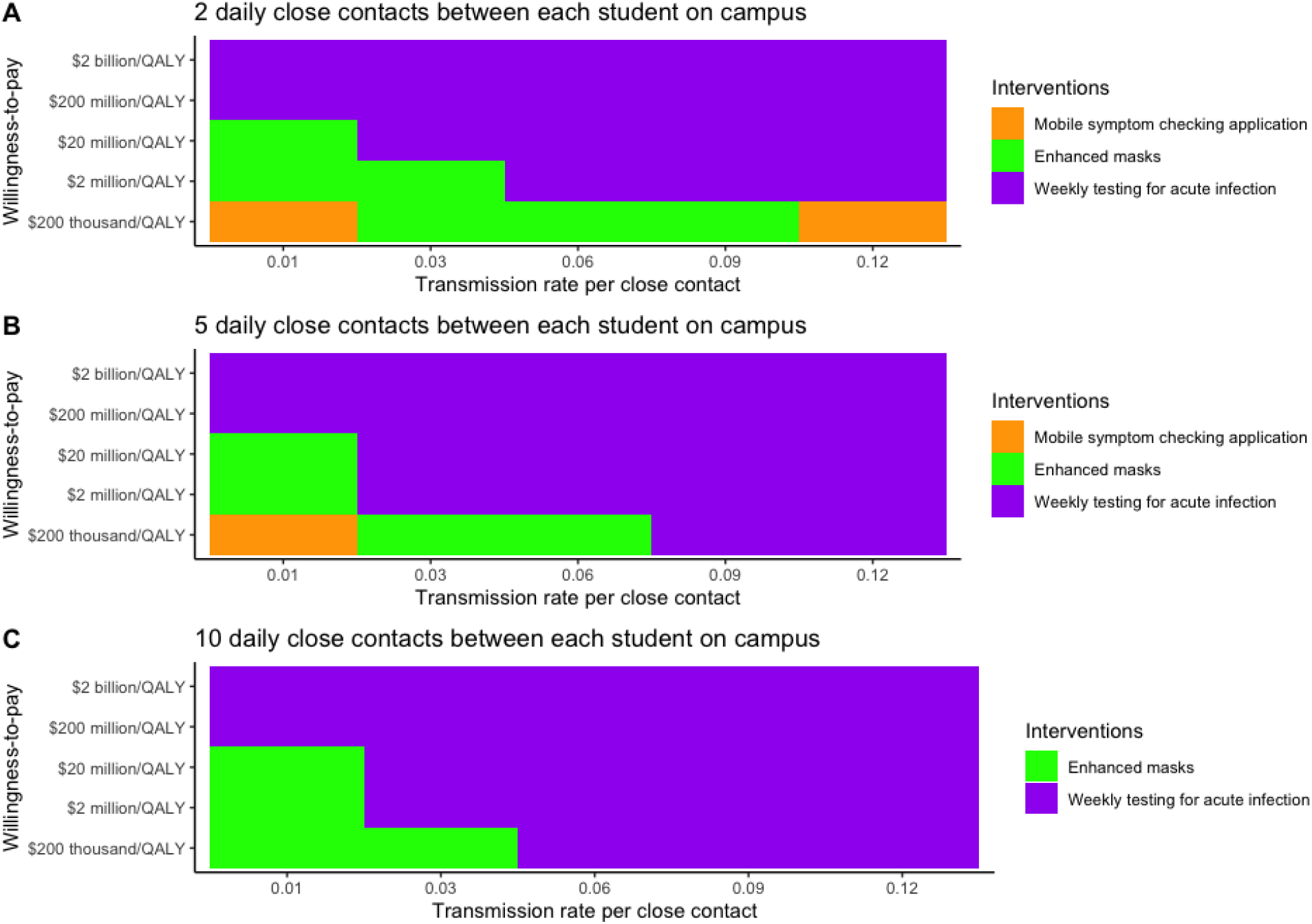
Three-way sensitivity analysis examining the relationship between the number of close contacts between students on campus, the transmission rate per close student contact, and willingness-to-pay for the top 3 intervention strategies at a 2% prevalence rate of actively infectious cases in the community.

At a willingness-to-pay threshold of $200,000/QALY gained and a prevalence ranges of 0.3-0.5%, MERV-13 ventilation upgrades would only be cost-effective were fine aerosol transmission to account for >90% of all campus COVID-19 transmission. For far-UVC lighting, the threshold for fine aerosol transmission is >86%. As the proportion of students wearing masks on campus declines, the ICER of both of these systems becomes more favorable.

Including or excluding faculty and staff over age 70 years had no substantial impact under any scenario because of their relatively small numbers. Across all prevalence rates, increasing the chance of a super-spreader event reduced the cost-effectiveness of interventions because such events lead to early closure of the campus. Additional sensitivity analyses are available in the Online Appendix.

## Discussion

Our model shows that the value of most of the interventions is dependent on the prevalence of actively infectious cases of COVID-19 in the community surrounding the university. When case rates in the surrounding community are low (e.g., as in New York in July, 2020) the most value will be realized from simply using a symptom checking application. However, variables such as mask use and the number of contacts between affiliates are highly influential, and readers are strongly encouraged to change the model inputs to suit their particular university characteristics using the online version of the model. ^7^

While a QALY is typically valued at $100,000-$200,000, larger values are placed on nuclear or aviation regulations—over $1 million/QALY gained—because of the higher perceived threat amongst the public.^22-24^ COVID-19 may merit higher investments because of larger perceived threat of the pandemic.

A recent study by Paltiel and colleagues recommended testing for SARS-CoV-2 when the prevalence is 0.2%. ^25^ We find that gateway or weekly testing would be costly at this prevalence rate at Columbia University. These different conclusions can be attributed to differences in our: 1) infection fatality rate, 2) risk of transmission on campus, 3) the number of close contacts/student, and 4) outcome measure (we used cost/QALY gained as an outcome measure while they used cost/case detected). **Figure 1** shows as the number of close contacts between students increases and the rate of transmission increases, so too does the value of weekly testing. If students return to multi-generational households, we would expect to find similar results.

Our infection fatality rate (0.02% for students and 0.18% for staff/faculty, **Table 1**) is smaller than the average rate for the U.S. (0.5%)^14,25^ because the population of both the students and staff/faculty is younger than the general population. Users of our online model should be careful to define risks specific to their university setting.

Many universities plan to standardize the masks that students wear, such that their fit and filtration are superior to what students would choose to purchase on their own. ^26,27^ For example, Columbia University will provide two $4 2-ply masks to each student. ^6^ Such masks would be cost-saving at community prevalence rates of infectious cases above 0.3% at Columbia University.

Fever is a common symptom in the fall. ^28^ Even though thermal monitoring will detect few cases of COVID-19, it will send a proportion of university affiliates home, thereby increasing social distancing and the prevalence of people on campus with a potentially infectious influenza-like illness. This strategy nevertheless comes at a low value under most circumstances.

Likewise, installations of high efficiency MERV-13 HVAC systems or far UVC light are not likely to be worthy of investment unless aerosol transmission accounts for the vast majority of COVID-19 cases on campus. However, the value of these systems may rise in areas where students are less likely to wear masks, such as in student housing. Additionally, while we find that far UVC light comes at a better value than HVAC systems, it is a new technology and its safety has not been extensively tested in humans. ^29^ It also has the value of reducing fomite transmission, a factor for which we did not account.

We assumed that super-spreader events would require the university to close because they may create a “perfect storm” of events in which staff are quarantined even as multiple areas of the campus must be closed for cleaning and contact tracing. When the university is closed early, the money spent on any interventions goes to waste, and large indirect costs associated with online-only instruction are incurred. Therefore, any intervention to reduce the possibility of students attending mass events, particularly those that require talking loudly and lowering masks to eat or drink, should be prioritized.

We did not examine approaches that are intuitively cost-effective. For example, influenza vaccination has been shown to save money and lives. ^30^ Likewise, the “cohort” model confines groups of students to 20-40 who take all the same courses and live in the same housing. ^31^ In theory, the cohort model could largely prevent transmission of infection, thereby vastly reducing the size of each student’s social network.

The major limitation of our analysis is the considerable uncertainty in parameter estimates. For example, estimates of infection fatality rates can quadruple when hospitals are overwhelmed with cases.^14,17^ However, the model is generally robust to different parameter inputs and assumptions.

Another limitation is that universities vary considerably with respect to sociodemographic composition and risk-taking among students. Additionally, the standard gamble exercises we used were administered to graduate students of public health, who may be more risk adverse than average students. We account for differences in student risk preferences by varying the number of assumed contacts between students, both on and off campus in sensitivity analyses.

Our findings do not apply to universities in which a large number of students commute to and from multi-generational households.

## Conclusions

Individual universities are likely to find very different value in the interventions we present. Most universities will find value in strategies to reduce the transmission of COVID19 only if the prevalence of infectious cases in the community is high enough for the interventions to impact transmission of SARS-CoV-2, but not so high that the university will need to close early.

## Data Availability

All data and the model code are available under a GNU 3.0 license. Feel free to use the model. If you have any bugs to report, please contact the authors.

https://www.publichealth.columbia.edu/academics/departments/health-policy-and-management/openup-model

## Acknowledgements

We would like to acknowledge the help and contributions of Wafaa El-Sadr, Melanie Bernitz, Steven Shea, Wan Yang, Jeffery Shamen, and the Public Health Committee for Reopening Columbia University.

